# Incidence rate, risk factors and patient reported outcome in patients with a dislocation following hip hemiarthroplasty after acute femoral neck fracture; a scoping review

**DOI:** 10.1101/2024.04.29.24306544

**Authors:** Susanne C. Faurholt Närhi, Louise I. E. Ø. Rasmussen, Søren Overgaard, Bjarke L. Viberg, Lars L. Hermansen

## Abstract

**Objective:** The objective of this scoping review of the literature is to find the incidence rate and risk factors for dislocation of hip hemiarthroplasties (HAs) after acute femoral neck fractures (FNFs). Additionally we aim to determine the subjectively reported experience and/or Patient Reported Outcomes (PROs) minimum six months after a dislocation of a hip HA after acute FNF.

**Introduction:** The existing literature suggests a dislocation rate of 1 – 12%(1-8), and we aim to evaluate the dislocation rates reported in the literature, and explain the differences in the reportings. Some of the suggested risk factors ranging from surgical approach(1, 5, 6), cognitive impairment(2, 5-8), prosthesis type(4) and uncemented vs cemented implant(3). There are no studies summarizing all risk factors for dislocating the hip HA. The patient’s subjective experience after dislocation of a hip HA after acute FNF is not well known. The literature lacks direct information of the patients’ subjective experience after dislocation of the hip HA after acute FNF.

**Inclusion criteria:** Published articles on the incidence of patients with HA, who develop dislocation. Risk factors for dislocation and patient reported outcomes after reposition of dislocation. Register studies, clinical prospective studies and case-control studies will be included. The lower limit for inclusion of a risk factor will be minimum 5 studies that have minimum 10 patients with dislocation(s).

**Methods:** This scoping review will be conducted in accordance with the JBI methodology for scoping reviews(9). We will develop a full search strategy for Embase, MEDLINE, PubMed and Cochrane Library. Studies published in English, Swedish, Danish and Norwegian will be included. Studies with other languages will be considered if an appropriate translator is available. The three research questions will be analyzed separately and reported narratively. Despite this being a scoping review, we shall include some risk of bias elements in the analysis.

## Introduction

In Denmark, about 7.000 patients are admitted with an acute femoral neck fracture (FNF) every year(10). Depending on the fracture morphology, treatment choice differs. Treatment with a hip hemiarthroplasty (HA) is the treatment choice for the majority of displaced fractures (Garden type III-IV)(11). Dislocation of the HA seems to be a relatively common complication, and we know that it is a cause of considerable pain, and we believe it limits the patient’s daily living after they have experienced a dislocation once. The treatment for a dislocated HA is usually closed reduction, while the patient is under a short-lasting general anesthesia.

### Dislocation rate

Unfortunately, dislocation of the HA lacks an ICD-10 diagnosis code, why it is difficult to find the precise number of people affected. The existing literature is unclear of the dislocation rates of the hip HA, ranging from 1 – 12%(1-8). Many studies describing prevalence, incidence or incidence rate base their dislocation rate on patients treated at their facility or insurance data. Hence, there is a high risk of loss of follow-up and/or bias. We aim to evaluate the dislocation rates reported in the literature, and explain the differences in the reportings.

### Risk factors for dislocation

Knowledge of reasons for dislocation of the hip HA would help surgeons choose, or opt out, on the implant when the patient is deemed too high-risk for dislocation, or we might be able to warn the patients’ of the possible risks and push them to be more careful. Thus, there are several studies suggesting some risk factors for dislocation of the hip HA, such as surgical approach(1, 5, 6), cognitive impairment(2, 5-8), prosthesis type(4) and uncemented vs cemented implant(3). However, such studies often contradict each other, and there is a lack of conclusive evidence and the hierarchy of the proposed risk factors are unknown. Furthermore, there are no studies summarizing all risk factors for dislocating the hip HA.

### Long-term quality of life following dislocation of the hip HA

After the patient has experienced a dislocation of the hip HA, and the hip HA has been reduced at the hospital, the patient is discharged from the hospital and there is no follow-up of the patient at all. Therefore, we, as surgeons, do not know how the dislocation affects the patient’s daily living. We do not know if the patient can live their life as they used to. The literature lacks direct information of the patients’ subjective experience of the quality of life after dislocation of the hip HA after acute FNF.

A preliminary search of PubMed, MEDLINE, Cochrane Library and Embase indicates that there are some systematic reviews on the incidence rate, however, the reviews are not focused on only dislocation of the hip HA. We found two systematic reviews of risk factors, and no systematic- or scoping reviews on these three topics.

One of the two systematic reviews focusing on a summation of risk factors of dislocating the hip HA after FNF(12, 13) were not focused on only risk factors for dislocating the hip HA, but also included total hip arthroplasties (THAs). However, we found no reviews at all on long-term PROMs or the subjective experience following dislocation of the hip HA. We see a need to summarize these three topics.

The objectives of this scoping review are:

i. To identify the the incidence rate
ii. Obtain an overview of risk factors for dislocation of hip HAs after acute FNFs.
iii. Gain an understanding of the long-term quality of life in patients experience/the subjectively reported experience and/or PROs minimum 6 months after a dislocation of a hip HA after acute FNF.

This knowledge will be of great value for the clinician, who will be well equipped to correctly inform the patient of the frequency and risk factors of dislocation, and when HA patients are faced with dislocation need information about what they can expect to experience after the dislocation.

## Review question

### Our question of interest

“What is the incidence rate, risk factors and long-term patient reported outcomes for dislocating a hip hemiarthroplasty after acute femoral neck fracture?”

## Eligibility criteria

Studies will be included in the review if the following criteria are fulfilled:

Study designs: register studies, clinical prospective studies and case-control studies. The lower limit for inclusion of a risk factor will be minimum 5 studies that have minimum 10 patients with dislocation(s). We have based the lower limit for inclusion of a risk factor on expertise within the research group, as to set a lower limit on when to accept a potential risk factor as a risk factor.

### Participants

Acute femoral neck fracture as primary diagnosis. HA regardless of surgical approach and type of components.

Follow-up of at least 3 months of follow-up is required. A maximum of follow-up is set for 5 years after last episode of dislocation regarding PRO as outcome.

No “prior to revision surgery” studies are included (selected patients).

### Concept

Incidence of patients with HA, who develop dislocation. Risk factors for dislocation and patient reported outcomes after reposition of dislocation.

### Context

No specific context as long as the above is fulfilled. We will include publications from all parts of the world, including all races, genders and ages.

### Types of Sources

- Objective i) randomized controlled trials, non-randomized controlled trials, before and after studies and analytical observational studies including prospective and retrospective cohort studies, case-control studies and analytical cross-sectional studies.
- Objective ii) randomized controlled trials, non-randomized controlled trials, before and after studies, analytical observational studies including prospective and retrospective cohort studies, case-control studies and analytical cross-sectional studies, descriptive observational study designs including case series, and descriptive cross-sectional.
- Objectve iii) prospective and retrospective cohort studies, qualitative studies including, but not limited to, designs such as phenomenology and qualitative description.

## Methods

We have chosen a scoping approach in this study, mainly because we do not consider it necessary to do a full risk of bias analysis on each topic of this scoping review. However, we aim to assess the quality of the included studies in some degree, see section “Data analysis and presentation”. This scoping review will be conducted in accordance with the JBI methodology for scoping reviews(9).

### Search strategy

The search strategy will aim to locate both published and unpublished studies. An initial limited search of Embase, MEDLINE, PubMed and Cochrane Library was undertaken to identify articles on the topic. The text words contained in the titles and abstracts of relevant articles, and the index terms used to describe the articles were used to develop a full search strategy for Embase, MEDLINE, PubMed and Cochrane Library (see Appendix I). The search strategy, including all identified keywords and index terms, will be adapted for each included database and/or information source. The reference list of all included sources of evidence will be screened for additional studies.

Studies published in English, Swedish, Danish and Norwegian will be included. Studies with other languages will be considered if an appropriate translator is available. Studies will be included without time limitation, but we will consider the study’s date in the analysis phase, since some prosthesis types may be outdated.

The databases to be searched include PubMed, MEDLINE, Embase and Cochrane Library. Sources of unpublished studies/ gray literature to be searched include trial registers in Cochrane Library.

### Study/Source of Evidence selection

Following the search, all identified citations will be collated and uploaded into ©2024 Covidence and the duplicates will be removed. Following a pilot test, titles and abstracts will then be screened by two independent reviewers for assessment against the inclusion criteria for the review. Potentially relevant sources will be retrieved in full and their citation details will be in full in Covidence. The full text of selected citations will be assessed in detail against the inclusion criteria by two independent reviewers. Reasons for exclusion of sources of evidence at full text that do not meet the inclusion criteria will be recorded and reported in the scoping review. Any disagreements that arise between the reviewers at each stage of the selection process will be resolved through discussion, or with an additional reviewer. The results of the search and the study inclusion process will be reported in full in the final scoping review and presented in a Preferred Reporting Items for Systematic Reviews and Meta-analyses extension for scoping review (PRISMA-ScR) flow diagram(14, 15).

### Data Extraction

Data will be extracted from papers included in the scoping review by two independent reviewers by hand, using Excel. The data extracted will include specific details about the participants, concept, context, study methods and key findings relevant to the review question.

Before conducting the review, we will create a standardized data extraction form, which will include guidance of what will be extracted, see appendix III.

### Data Analysis and Presentation

The data will be presented graphically or in a tabular form, with a narrative summary accompanying the graphs or tables. In the narrative summary, a description of the relation between the results and objective of this study will be presented.

For objective i), we expect to report: 1. Study design (single center vs multi center), 2. Time factor (weeks, months, year(s)) and 3. Completeness (is the follow-up sufficient, high drop out?). This will be the quality assessment of the study and will be presented in a separate table.

For objective ii) we aim to report whether the risk factors had been validated and/or is present in five or more publications. It also includes reports with negative findings. This will be reported in a separate table.

Objective iii) The subjective experience after dislocation will be assessed according to completeness (which patients agreed to be included in the studies?), and will be reported either graphically or in a table.

These results will be taken into account when answering the research questions.

## Data Availability

All data is avaliable via PubMed, Embase, Medline and Cochrane Library.

## Acknowledgements

Mette Brandt Eriksen, research librarian, associate professor MSc, PhD. for the patience, lessons and great help and instructions with the literature search.

Peter Everfeldt, medical librarian, for the help with getting access to non-open access publications.

Professor Emerita Charlotte Leboeuf-Yde for assistance during the project.

This protocol was drafted from the JBI Scoping Review Network’s protocol template(16).

## Appendices

### Appendix I: Search strategy

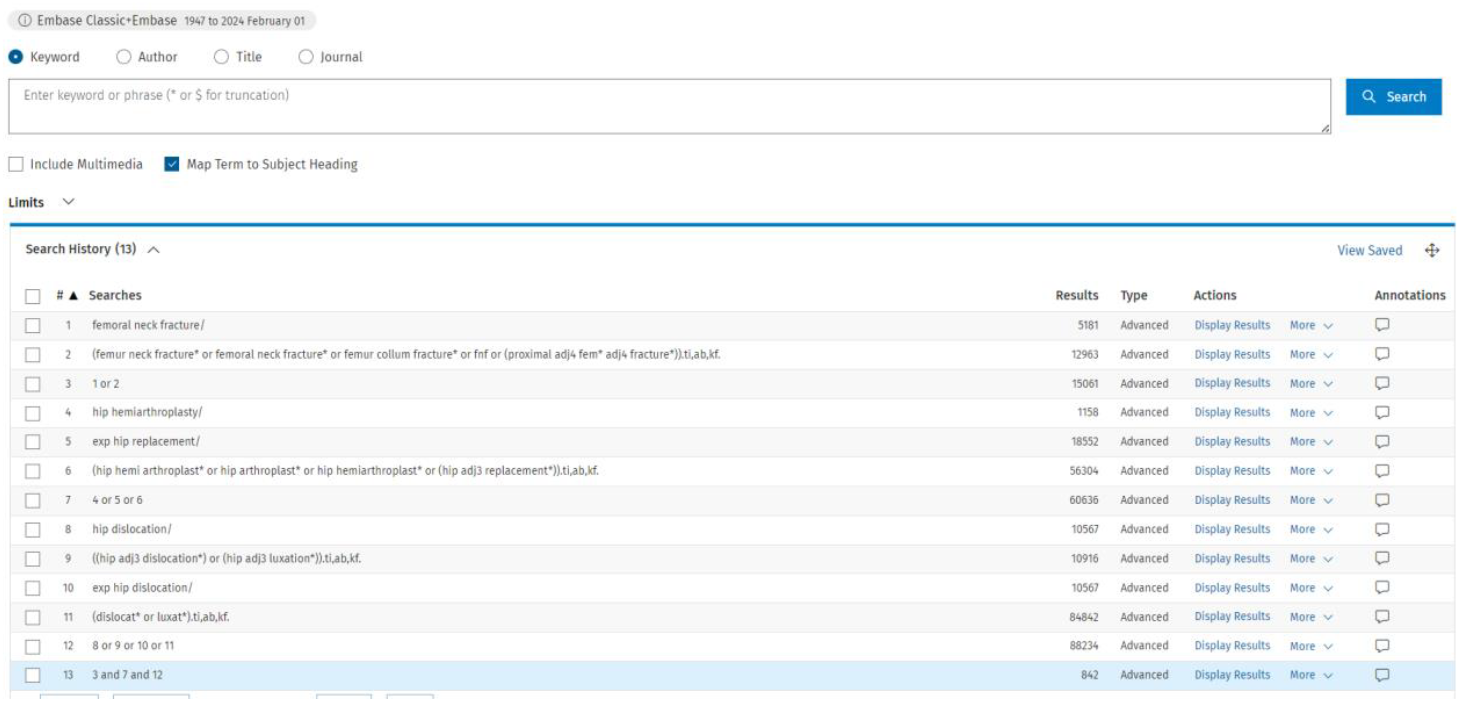

### Appendix II: Data extraction instrument

Covidence, Excel

### Appendix III

#### Data Extraction Sheet/Descriptive table

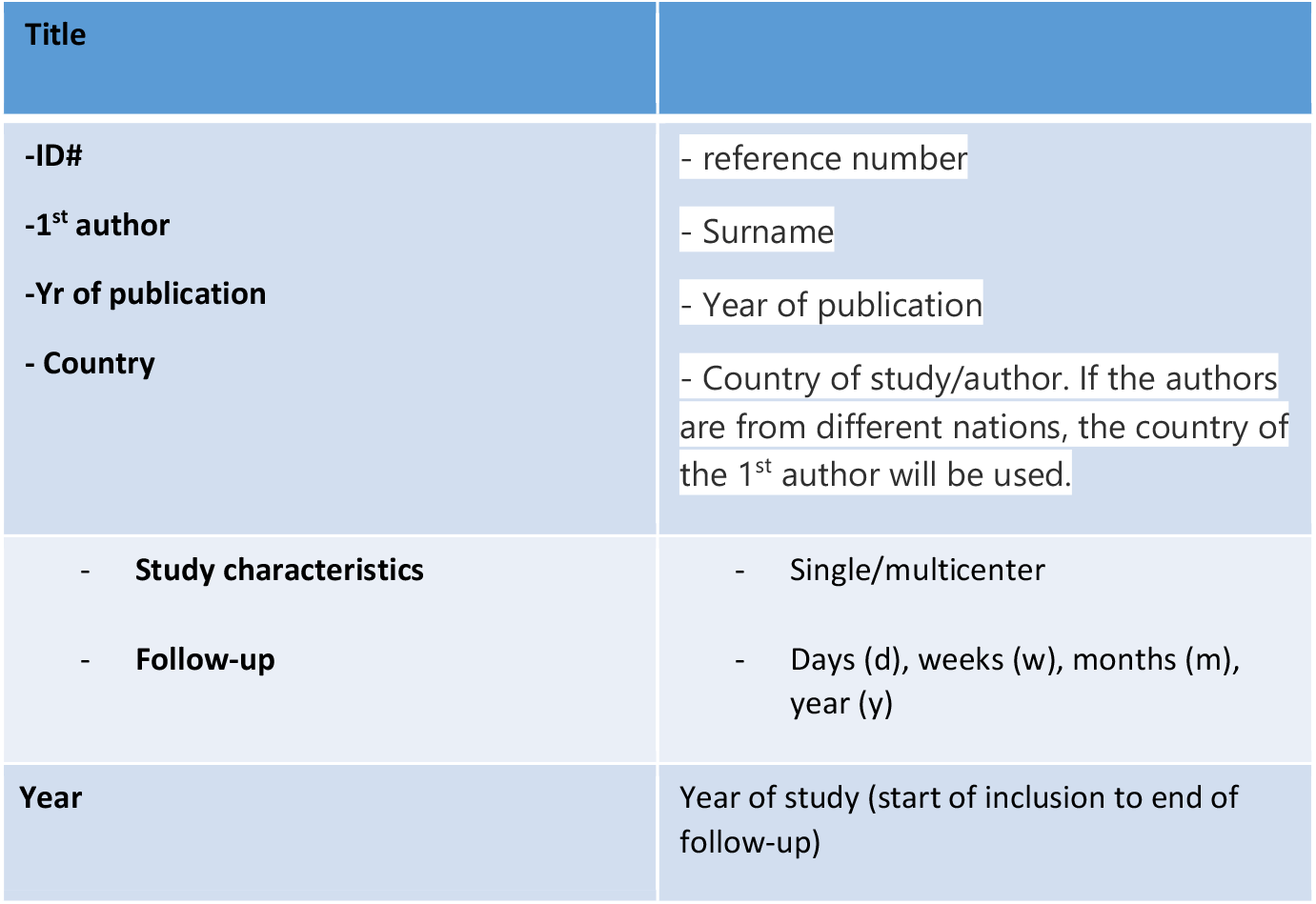
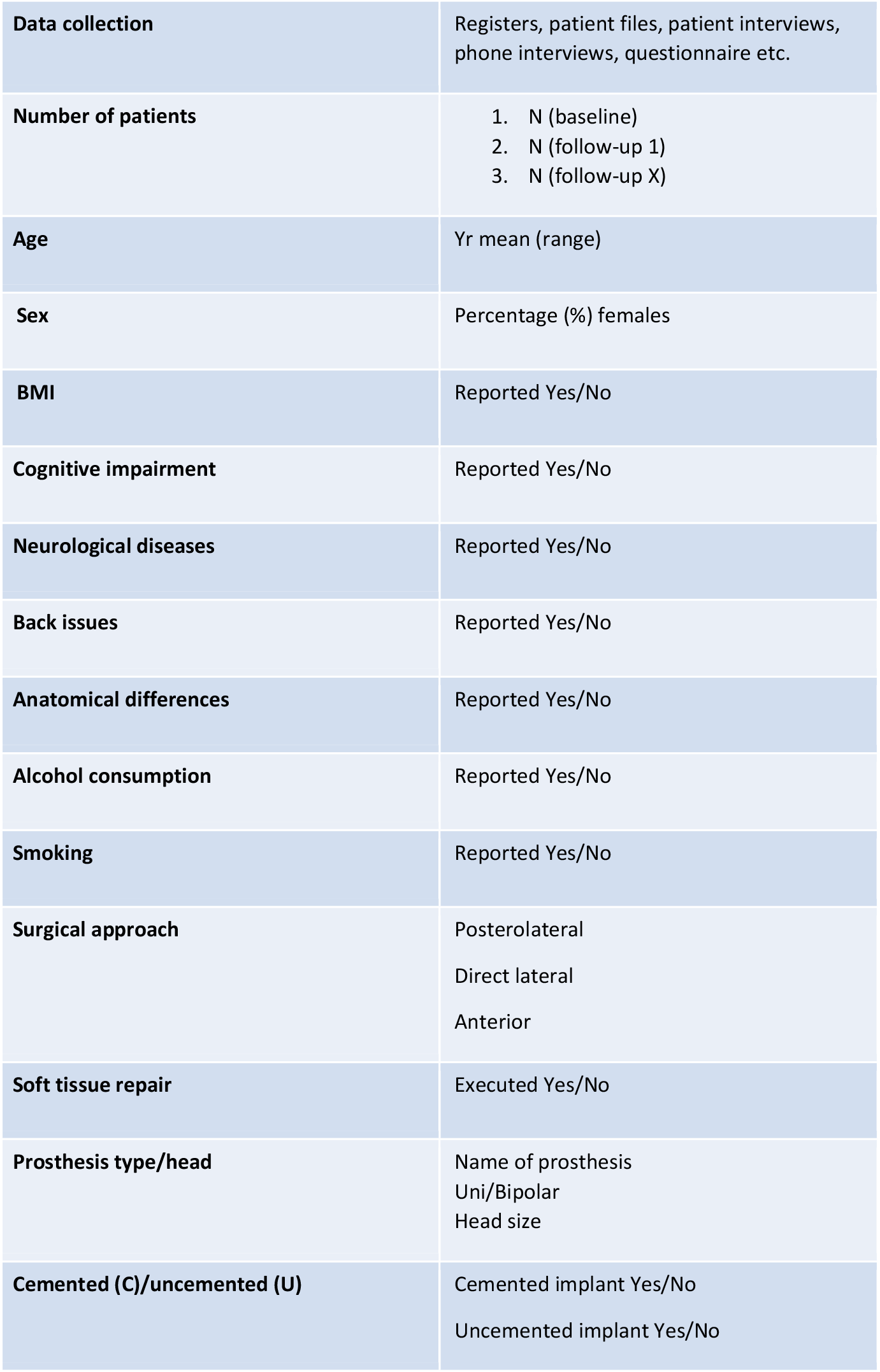
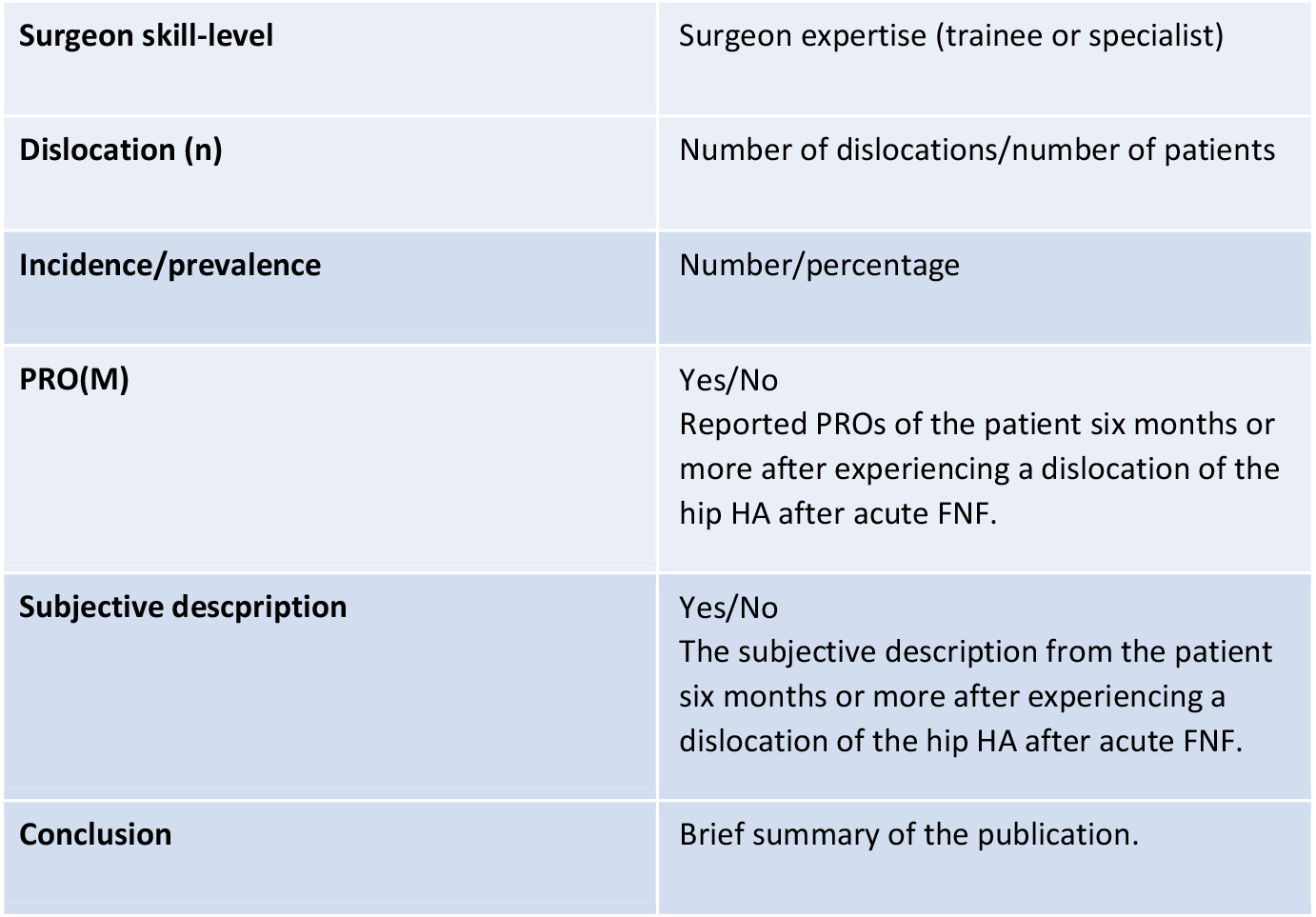

| Title |  |
| --- | --- |
| -ID#<br>-1<sup>st</sup> author<br>-Yr of publication<br>- Country | - reference number<br>- Surname<br>- Year of publication<br>- Country of study/author. If the authors are from different nations, the country of the 1<sup>st</sup> author will be used. |
| - Study characteristics<br>- Follow-up | - Single/multicenter<br>- Days (d), weeks (w), months (m), year (y) |
| Year | Year of study (start of inclusion to end of follow-up) |
| **Data collection** | Registers, patient files, patient interviews, phone interviews, questionnaire etc. |
| **Number of patients** | 1. N (baseline)<br>2. N (follow-up 1)<br>3. N (follow-up X) |
| **Age** | Yr mean (range) |
| **Sex** | Percentage (%) females |
| **BMI** | Reported Yes/No |
| **Cognitive impairment** | Reported Yes/No |
| **Neurological diseases** | Reported Yes/No |
| **Back issues** | Reported Yes/No |
| **Anatomical differences** | Reported Yes/No |
| **Alcohol consumption** | Reported Yes/No |
| **Smoking** | Reported Yes/No |
| **Surgical approach** | Posterolateral<br>Direct lateral<br>Anterior |
| **Soft tissue repair** | Executed Yes/No |
| **Prosthesis type/head** | Name of prosthesis<br>Uni/Bipolar<br>Head size |
| **Cemented (C)/uncemented (U)** | Cemented implant Yes/No<br>Uncemented implant Yes/No |
| **Surgeon skill-level** | Surgeon expertise (trainee or specialist) |
| **Dislocation (n)** | Number of dislocations/number of patients |
| **Incidence/prevalence** | Number/percentage |
| **PRO(M)** | Yes/No<br>Reported PROs of the patient six months or more after experiencing a dislocation of the hip HA after acute FNF. |
| **Subjective description** | Yes/No<br>The subjective description from the patient six months or more after experiencing a dislocation of the hip HA after acute FNF. |
| **Conclusion** | Brief summary of the publication. |

#### Report

##### Dislocation rate

|  |  |
| --- | --- |
| **-ID#**<br>**-1<sup>st</sup> author**<br>**-Yr of publication** |  |
| **Study design** | Single- or multicentre, register study |
| **Follow-up rate** | Duration of follow-up (Weeks (w), months (m), year (y)) |
| **Completeness** | Nominator (Number of patients with dislocation) /Denominator (Number of patients with HA after FNF) |

##### Risk factors

|  |  |
| --- | --- |
| **-ID#**<br>**-1<sup>st</sup> author**<br>**-Yr of publication** |  |
| **Risk factor\* 1** | Validated? |
| **Risk factor 2** | Validated? |
| **Risk factor xx** |  |
| **Completeness** | Sufficient follow-up/high drop out |

\*A risk factor will only be reported if five or more studies report that risk factor (positive and negative findings), with at least 10 patients with dislocated HAs.

##### Subjective experience of dislocation/PROs

|  |  |
| --- | --- |
| **-ID#**<br>**-1<sup>st</sup> author**<br>**-Yr of publication** |  |
| **Invited patients** | Number (n) |
| **Accepted** | Number (%) |
| **Participated** | Number (%) |
| **Completed the study** | Number (%) |
| **Follow-up rate** | Duration of follow-up (Weeks (w), months (m), year (y)) |

## References

1. Viberg B, Kristensen EQ, Gaarsdal T, Petersen CD, Jensen TG, Overgaard S, et al. A piriformis-preserving posterior approach reduces dislocation rate of the hemiarthroplasty in patients with femoral neck fracture. Injury. 2023.

2. Awadallah M, Blanco J, Ong J, Kumar N, Rajata P, Parker M. Is there a higher risk of dislocation of hip hemiarthroplasty in patients with neuromuscular conditions? A clinical study of 3827 patients. Injury. 2022;53(2):631–3.

3. Jameson SS, Jensen CD, Elson DW, Johnson A, Nachtsheim C, Rangan A, et al. Cemented versus cementless hemiarthroplasty for intracapsular neck of femur fracture--a comparison of 60,848 matched patients using national data. Injury. 2013;44(6):730–4.

4. Joanroy R, Stork-Hansen J, Rotwitt L, Viberg B. Cemented hemiarthroplasty for femoral neck fracture patients: collarless, polished tapered stem (CPT) versus anatomic matte stem (Lubinus SP2). Eur J Orthop Surg Traumatol. 2021;31(5):855–60.

5. Jobory A, Karrholm J, Hansson S, Akesson K, Rogmark C. Dislocation of hemiarthroplasty after hip fracture is common and the risk is increased with posterior approach: result from a national cohort of 25,678 individuals in the Swedish Hip Arthroplasty Register. Acta Orthop. 2021;92(4):413–8.

6. Kristoffersen MH, Dybvik E, Steihaug OM, Kristensen TB, Engesaeter LB, Ranhoff AH, et al. Cognitive impairment influences the risk of reoperation after hip fracture surgery: results of 87,573 operations reported to the Norwegian Hip Fracture Register. Acta Orthop. 2020;91(2):146–51.

7. Page BJ, Parsons MS, Lee JH, Dennison JG, Hammonds KP, Brennan KL, et al. Surgical Approach and Dislocation Risk After Hemiarthroplasty in Geriatric Patients With Femoral Neck Fracture With and Without Cognitive Impairments-Does Cognitive Impairment Influence Dislocation Risk? J Orthop Trauma. 2023;37(9):450–5.

8. Olesen BA, Narhi SF, Jensen TG, Overgaard S, Palm H, Sorensen MS. Incidence of dislocation and associated risk factors in patients with a femoral neck fracture operated with an uncemented hemiarthroplasty. BMC Musculoskelet Disord. 2024;25(1):119.

9. Peters MDJ GC, McInerney P, Munn Z, Tricco AC, Khalil, H. Chapter 11: Scoping Reviews (2020 version) JBI Manual for Evidence Synthesis: JBI; 2020; 2020 [Available from: https://synthesismanual.jbi.global.

10. Jantzen C, Madsen CM, Lauritzen JB, Jorgensen HL. Temporal trends in hip fracture incidence, mortality, and morbidity in Denmark from 1999 to 2012. Acta Orthop. 2018;89(2):170–6.

11. Palm H, Krasheninnikoff M, Holck K, Lemser T, Foss NB, Jacobsen S, et al. A new algorithm for hip fracture surgery. Reoperation rate reduced from 18 % to 12 % in 2,000 consecutive patients followed for 1 year. Acta Orthop. 2012;83(1):26–30.

12. Wang B, Liu H, Zhu Y, Yan L, Li JJ, Zhao B. Risk Factors with Multilevel Evidence for Dislocation in Patients with Femoral Neck Fractures After Hip Hemiarthroplasty: A Systematic Review. Indian J Orthop. 2020;54(6):795–804.

13. Hernigou P, Barbier O, Chenaie P. Hip arthroplasty dislocation risk calculator: evaluation of one million primary implants and twenty-five thousand dislocations with deep learning artificial intelligence in a systematic review of reviews. Int Orthop. 2023;47(2):557–71.

14. Tricco AC, Lillie E, Zarin W, O’Brien KK, Colquhoun H, Levac D, et al. PRISMA Extension for Scoping Reviews (PRISMA-ScR): Checklist and Explanation. Ann Intern Med. 2018;169(7):467–73.

15. McGowan J, Straus S, Moher D, Langlois EV, O’Brien KK, Horsley T, et al. Reporting scoping reviews-PRISMA ScR extension. J Clin Epidemiol. 2020;123:177–9.

16. Network TJSR. JBI_Protocol_Template_Scoping_Reviews_2024 2024 [Available from: https://jbi.global/scoping-review-network/resources.

